# Trends in second-line glucose-lowering therapy initiations and treatment outcomes across age and frailty groups in people with type 2 diabetes: UK population-based study, 2019-2024

**DOI:** 10.64898/2026.01.07.26343532

**Authors:** MM Dinsdale, KG Young, P Cardoso, LM Güdemann, TT Jansz, AP McGovern, AG Jones, ER Pearson, AT Hattersley, TJ McKinley, BM Shields, JM Dennis

## Abstract

**Objective:** To assess contemporary UK population-level trends in second-line glucose-lowering therapy initiation and explore treatment outcome trends across age and frailty groups in people with type 2 diabetes (T2D).

**Methods:** We studied 117,064 adults with T2D who initiated major second-line glucose-lowering drug classes after metformin between 2019–2024 in population-representative UK data (Clinical Practice Research Datalink). Outcomes were assessed in three groups: age ≤70 years (n=84,589 [72%]), non-frail >70 years (n=18,933 [16%]), and frail >70 years (n=13,524 [12%]), with frailty specified as an electronic frailty index ≥ 0.24. Annual prescribing trends were calculated as the proportion of each drug class among all second-line initiations. We evaluated 12-month changes in HbA_1c_, weight, treatment discontinuation, and complications, including composite severe diabetes-related complications, heart failure and kidney failure. Trends were estimated using multivariable regression models adjusted for demographic and clinical characteristics.

**Results:** Use of SGLT2-inhibitors increased from 26% of overall second-line initiations in 2019 to 63% in 2024, with the greatest increase among frail adults >70 (6% in 2019 vs 60% in 2024). DPP4-inhibitor use declined substantially over the same period (44% 2019 vs 16% in 2024). In adults aged ≤70 years, mean 12-month HbA_1c_ response improved from -11.3 mmol/mol (95%CI -11.6 to -11.0) in 2019 to -12.9 mmol/mol (95%CI -13.2 to -12.6) in 2023, and mean weight loss improved from -1.7 kg (95%CI -1.8 to -1.5) to -2.8 kg (95%CI -2.9 to -2.6). Treatment discontinuation within 12 months remained stable (30.2% [95%CI 29.4 to 31.0] in 2019 vs 30.7% [95%CI 30.0 to 31.4] in 2022). There was limited evidence of change in short-term complications over 2019-2022: rates (per 1,000 person-years) of severe diabetes-related complications were 9.9 (95%CI 8.1 to 11.7) vs 8.6 (95%CI 6.9 to 10.3), heart failure 2.0 (95%CI 1.1 to 2.9) vs 2.7 (95%CI 1.8 to 3.7), and kidney failure 2.1 (95%CI 1.1 to 3.0) vs 1.2 (95%CI 0.5 to 1.8). All trends were consistent in adults >70, including those with frailty.

**Conclusion:** Use SGLT2-inhibitors in people with T2D has rapidly increased in the UK over 2019–2024 to represent the majority of second-line initiations, with a 10-fold increase in initiations amongst older adults with frailty. Over the same period, population-level benefits for HbA_1c_ and weight were modest, with no evidence of changes in risk of short-term discontinuation or diabetes complications. Findings support the careful use of SGLT2i across the wider population with type 2 diabetes including frail older adults.

## INTRODUCTION

Type 2 diabetes affects over 700 million people,^1^ and most individuals will require glucose-lowering therapy in their lifetime to achieve and maintain glycaemic control. Metformin is almost universally recommended as first-line treatment,^2^ but for many people there is a lack of guidance on which second-line therapy to prescribe after metformin and recommendations differ between national and international guidelines.^3,4^ In practice, this lack of clarity means treatment decisions are often shaped by clinician and patient preference, resulting in marked variability by geography and patient characteristics.^5,6 7 8 9^

Newer T2D therapies such as SGLT2-inhibitors (SGLT2i) offer benefits beyond glycaemic control, including weight loss and cardiorenal protective effects.^10^ Reflecting this evidence, international and UK National Institute for Health and Care Excellence (NICE) treatment guidance now recommend SGLT2i independently of glycaemic control for people with cardiovascular or chronic kidney disease, and to consider them for those at high cardiovascular risk.^3,4^ Under NICE recommendations, over 90% of people with type 2 diabetes in the UK are now eligible for, or should be considered for, SGLT2i therapy.^11^ Despite this, recent studies across multiple countries have suggested uptake of SGLT2i remains lowest in those most likely to benefit, including those with cardiovascular or kidney disease and in older adults.^12, 13 14 15 16^ To date, however, no studies have examined UK type 2 diabetes prescribing trends beyond 2020 or recent trends in population level treatment outcomes after initiating therapy.^6^

We aimed to describe contemporary prescribing trends for second-line glucose-lowering therapies after metformin in the UK between 2019 and 2024, with a particular focus on frail older adults where SGLT2i have historically been prescribed with caution.^14^ We further evaluated trends in short-term clinical outcomes after second-line initiation, including glycaemic response, weight change, treatment discontinuation, and severe diabetes-related complications.

## METHODS

### Study design and participants

We conducted a population-based cohort analysis using data from the UK Clinical Practice Research Datalink (CPRD) Aurum, which contains UK-representative primary care data covering approximately 13% of the population in England.^17^ Primary care records were linked to hospital admission data (Hospital Episode Statistics [HES]), Office for National Statistics (ONS) death registrations and individual-level 2019 English Index of Multiple Deprivation (IMD).

We included adults diagnosed with type 2 diabetes who initiated second-line glucose-lowering therapy after initial treatment with metformin between 1 January 2019 and 1 March 2024. Cohort identification followed our previously published protocol^18^ (see https://github.com/Exeter-Diabetes/CPRD-Codelists for all codelists). HES data were available up to March 2023.

Second-line medications were categorised by major drug class into DPP4-inhibitors (DPP4i), GLP-1 receptor agonists (GLP-1RA), SGLT2i, sulphonylureas, and other (combined acarbose, glinides, thiazolidinediones and insulin, as prescribing numbers were low for each of these drug classes).

### Covariates

We extracted baseline participant characteristics at the time of second-line treatment initiation including age, sex, duration of diabetes, ethnicity (categorised according to major UK census categories: White, South Asian, Black, Mixed, Other), social deprivation (IMD quintile), BMI (kg/m^2^), weight (kg), HbA_1c_ (mmol/mol), cardiovascular disease (CVD: myocardial infarction, stroke, revascularisation, ischaemic heart disease, angina, peripheral arterial disease, transient ischaemic attack), chronic kidney disease (CKD) stages 3-5, heart failure, hypoglycaemia, hyperglycaemia, lower limb amputation, and severe retinopathy (vitreous haemorrhage, retinal photocoagulation).

### Age and frailty subgroups

Temporal prescribing trends and clinical outcomes were assessed within three subgroups defined by age and frailty status at second-line therapy initiation: age ≤70 years, non-frail >70 years and frail >70 years. Frailty was measured using the electronic frailty index (eFI),^19^ a validated cumulative deficit model based on the presence of 36 health deficits recorded in primary care. Each individual’s eFI score was calculated as the proportion of recorded deficits prior to second-line therapy initiation. Individuals were classified as frail (moderate/severe frailty, eFI > 0.24) or non-frail (fit/mild frailty, eFI ≤ 0.24).^20^

### Study outcomes

We examined calendar-year trends (2019 to 2024) in the initiation of second-line glucose-lowering therapies, as well as trends in short-term clinical outcomes after initiating therapy, including HbA_1c_ (glycaemic response), weight change, treatment discontinuation, and severe diabetes-related complications.

### Glycaemic response and weight change

We defined glycaemic response and weight change as the absolute change from baseline to 12 months following initiation of second-line therapy. Baseline HbA_1c_ was defined as the closest measurement within the 6 months prior to the drug start date. Baseline weight was defined as the closest measurement within the two years prior to the drug start date. HbA_1c_ and weight at 12-months were defined as the closest recorded value to 12 months post-treatment initiation, within a window of 9 to 15 months, with no addition or cessation of other glucose-lowering therapies, and continued prescription of the drug of interest. Outcomes were evaluated by calendar year of drug initiation over 2019 to 2023, allowing for a minimum of 12 months of follow-up within the study period to assess treatment effects.

### Treatment discontinuation

We defined treatment discontinuation as cessation of the drug of interest within 12 months of initiation. Patients were required to have 3 months of follow-up time after their last prescription to confirm that the drug was discontinued. We included patients who initiated therapy between 2019–2022, allowing sufficient follow up time to accrue (12 months for potential treatment duration and an additional 3 months of follow-up).

### Diabetes-related complications

Severe diabetes-related complications were assessed as a composite outcome, measured as the first occurrence of any event after drug initiation, defined according to recent UK Prospective Diabetes Study (UKPDS) criteria^21^: sudden death; death due to hyperglycaemia or hypoglycaemia; fatal or non-fatal myocardial infarction, angina, heart failure, stroke, or kidney failure; death from peripheral vascular disease; amputation; and severe retinopathy (vitreous haemorrhage or retinal photocoagulation). Events were identified using HES and death registry data (primary cause only), with additional capture of kidney failure from primary care codes.

We assessed incidence rates of heart failure and kidney failure following second-line therapy initiation separately, given the established benefits of SGLT2i for both endpoints.^22,23^ We also evaluated diabetic ketoacidosis (DKA) as a separate outcome, given the known risk associated with SGLT2i use.^24^

Complication outcomes were evaluated for patients initiating second-line therapy between January 2019 to December 2022 with follow-up until the earliest of: complication occurrence, discontinuation of second-line therapy, date of practice deregistration/death, the end of study period (March 2023; end of available follow-up in HES), or 1 year after treatment initiation.

## Statistical analysis

To evaluate prescribing trends for each drug class, we calculated the proportion of new second-line initiations for each drug class as a percentage of total second-line initiations for each calendar year.

### Glycaemic, weight, and treatment discontinuation outcomes

We evaluated non-linear time trends in glycaemic response and weight change for each calendar year using linear regression, with calendar year as a categorical covariate and an interaction between calendar year and age/frailty category. Estimates were adjusted for a standard covariate set comprising baseline HbA_1c_ (or baseline weight for weight outcome), age at therapy initiation, duration of diabetes, sex, ethnicity and deprivation. Missing ethnicity data were imputed using multiple imputation by chained equations.^25^ Treatment discontinuation was evaluated using logistic regression, adjusted for the same covariates.

### Severe diabetes-related complications

Adjusted incidence rates of severe diabetes-related complications, hospitalisation for heart failure and kidney failure were estimated using Poisson regression, with log follow-up time (in years) included as an offset term. The model was adjusted for the standard covariate set as well as prior history of the complication (composite, as previously defined). Due to the low number of events, only crude incidence rates were reported for DKA outcome.

To evaluate how clinical outcomes changed for each calendar year, independent of changes in the characteristics of patients initiating second-line therapy each year, we generated adjusted predictions from the fitted regression models. For each individual, we estimated the counterfactual outcome they would have if they had initiated therapy in each calendar year,^26^ holding all other covariates constant at their observed values. This produced a set of standardised predictions for each year to estimate expected outcomes, assuming no changes in the underlying population. We summarised overall temporal changes by comparing the adjusted predicted outcomes between the baseline year (2019) and the last year of follow-up (2022 or 2023, as appropriate).

### Impact of 2022 NICE guideline update

We further evaluated the impact on SGLT2i prescribing following the release of the 2022 UK NICE guidelines, which recommended SGLT2i use independent of glycaemic control for patients with CVD or CKD.^4^ Monthly SGLT2i prescribing rates were calculated as a proportion of all second-line initiations. We used an interrupted time series (ITS) autoregressive model to estimate pre-guideline trends, immediate changes following guideline introduction, and post-guideline trends.^27^ Counterfactual predictions were generated to estimate prescribing trends in the absence of the 2022 NICE guidelines.

All analyses were conducted using R (version 4.4.0), with key packages including mice (version 3.18.0) and tidyverse (version 2.0.0).^25, 28 29^

### Sensitivity analysis

To explore how prescribing trend changes related to patient characteristics beyond age and frailty, we further assessed trends across subgroups defined by sex, ethnicity, deprivation, and among patients with or without pre-existing CVD or CKD (stage 3-4).

To explore variations more thoroughly by age and frailty, we repeated all analyses with the frail >70 years group further stratified into those with moderate frailty (0.24 < eFI score ≤ 0.36) and those with severe frailty (eFI score > 0.36).

We repeated the glycaemic response and weight change analyses using change from baseline to 6 months (closest ±3 months, as for the definition of 12-month change) and similarly assessed treatment discontinuation within 6 months.

We ran further exploratory analysis of heart failure hospitalisations following second-line therapy initiation in individuals with pre-existing heart failure or cardiovascular disease. This aimed to assess whether short-term population-level benefits of cardioprotective therapies like SGLT2i were observed in those at highest baseline risk.

### Patient and public involvement

People with type 2 diabetes were involved in the MASTERMIND consortium and were key in identifying that better, more tailored, evidence was needed for the choice of glucose-lowering therapy. There was no patient or public involvement when conducting this specific study in terms of study design, analysis, interpretation or writing.

## RESULTS

### Study population

Between 2019 and 2024, 117,064 individuals with type 2 diabetes initiated second-line glucose-lowering therapy. Of these, 84,589 (72.3%) were aged ≤70 years, 18,933 (16.2%) were non-frail >70 years and 13,524 (11.5%) were frail >70 years. Table 1 shows the baseline characteristics of the study population by age and frailty status at second-line treatment initiation. Additional baseline characteristics by calendar year of therapy initiation and by frailty further stratified into moderate and severe frailty are provided in Supplementary Tables 1 and 2, respectively.

**Table 1.** Baseline characteristics of the study cohort at second-line treatment initiation.

| Variable | Age $\leq 70$ years | Non-frail $>70$ years | Frail $>70$ years |
| --- | --- | --- | --- |
| N (%) | 84,589 (72.3) | 18,933 (16.2) | 13,542 (11.5) |
| Age at therapy initiation (years) | 55.5 [9.6] | 76.2 [4.7] | 79.8 [6.0] |
| Sex (% Male) | 50,489 (59.7) | 11,625 (61.4) | 6967 (51.5) |
| Duration of diabetes (years) | 5.1 [4.3] | 8.4 [5.4] | 10.2 [5.9] |
| **Ethnicity (%)** |  |  |  |
| White | 64,398 (76.1) | 17,063 (90.1) | 12,218 (90.3) |
| South Asian | 11,959 (14.1) | 851 (4.5) | 796 (5.9) |
| Black | 4578 (5.4) | 391 (2.1) | 272 (2.0) |
| Mixed | 992 (1.2) | 103 (0.5) | 53 (0.4) |
| Other | 1596 (1.9) | 274 (1.4) | 89 (0.7) |
| Missing | 1066 (1.3) | 251 (1.3) | 96 (0.7) |
| **IMD quintile (%)** |  |  |  |
| 1 (least deprived) | 9810 (11.6) | 3584 (18.9) | 2165 (16.0) |
| 2 | 11,34 (13.4) | 3531 (18.6) | 2312 (17.1) |
| 3 | 12,630 (14.9) | 3083 (16.3) | 2162 (16.0) |
| 4 | 15,830 (18.7) | 2727 (14.4) | 2223 (16.4) |
| 5 (most deprived) | 18,463 (21.8) | 2338 (12.3) | 2200 (16.3) |
| Missing | 16,521 (19.6) | 3670 (19.5) | 2462 (18.2) |
| **Clinical features** |  |  |  |
| BMI (kg/m<sup>2</sup>) | 34.2 [7.7] | 30.3 [5.9] | 30.6 [6.5] |
| Weight (kg) | 98.5 [23.5] | 85.4 [17.8] | 83.8 [18.9] |
| HbA<sub>1c</sub> (mmol/mol) | 74.3 [20.3] | 68.7 [18.5] | 66.4 [19.9] |
| **Comorbidities (%)** |  |  |  |
| CVD<sup>a</sup> | 15,091 (17.8) | 5397 (28.5) | 8365 (61.9) |
| CKD stage 3-4 | 2709 (3.2) | 4191 (22.1) | 6254 (46.2) |
| CKD stage 5 | 258 (0.3) | 49 (0.3) | 83 (0.6) |
| Heart failure | 4485 (5.3) | 1607 (8.6) | 4863 (36.2) |
| Hyperglycaemia | 171 (0.2) | 22 (0.1) | 30 (0.2) |
| Hypoglycaemia | 52 (0.1) | 12 (0.1) | 15 (0.1) |
| Lower limb amputation | 179 (0.2) | 38 (0.2) | 80 (0.6) |
| Severe retinopathy<sup>b</sup> | 192 (0.2) | 109 (0.6) | 112 (0.8) |
| **eFI category<sup>c</sup> (%)** |  |  |  |
| Fit | 45,328 (53.6) | 5554 (29.3) | 0.0 |
| Mild | 30,701 (36.3) | 13,379 (70.6) | 0.0 |
| Moderate | 7227 (8.5) | 0.0 | 8961 (66.3) |
| Severe | 1333 (1.6) | 0.0 | 4563 (33.7) |

Values for continuous variables are given as mean [SD] and binary variables as n (%)
<sup>a</sup>CVD: myocardial infarction, stroke, revascularisation, ischaemic heart disease, angina, peripheral arterial disease, transient ischaemic attack
<sup>b</sup>Severe retinopathy: vitreous haemorrhage, retinal photocoagulation
<sup>c</sup>eFI category: fit ( $\text{eFI} \leq 0.12$ ); mild frailty ( $0.12 < \text{eFI} \leq 0.24$ ); moderate frailty ( $0.24 < \text{eFI} \leq 0.36$ ); severe frailty ( $\text{eFI} > 0.36$ ).

### Major increase in SGLT2i prescriptions across age and frailty subgroups over 2019-2024

We observed substantial changes in second-line prescribing between 2019 and 2024, with SGLT2i becoming the most initiated second-line therapy across all age and frailty subgroups. Overall, the proportion of SGLT2i prescriptions increased markedly from 26% of initiations in 2019 to 63% in 2024. This increase was most marked in those frail >70 years where the proportion of SGLT2i prescriptions increased 10-fold from 6% in 2019 to 60% in 2024 (Figure 1C). Similar trends were observed in those non-frail >70 years (12% in 2019 vs. 58% in 2024; Figure 1B) and in those aged ≤70 (30% in 2019 vs. 63% in 2024; Figure 1A).

**Figure 1.**
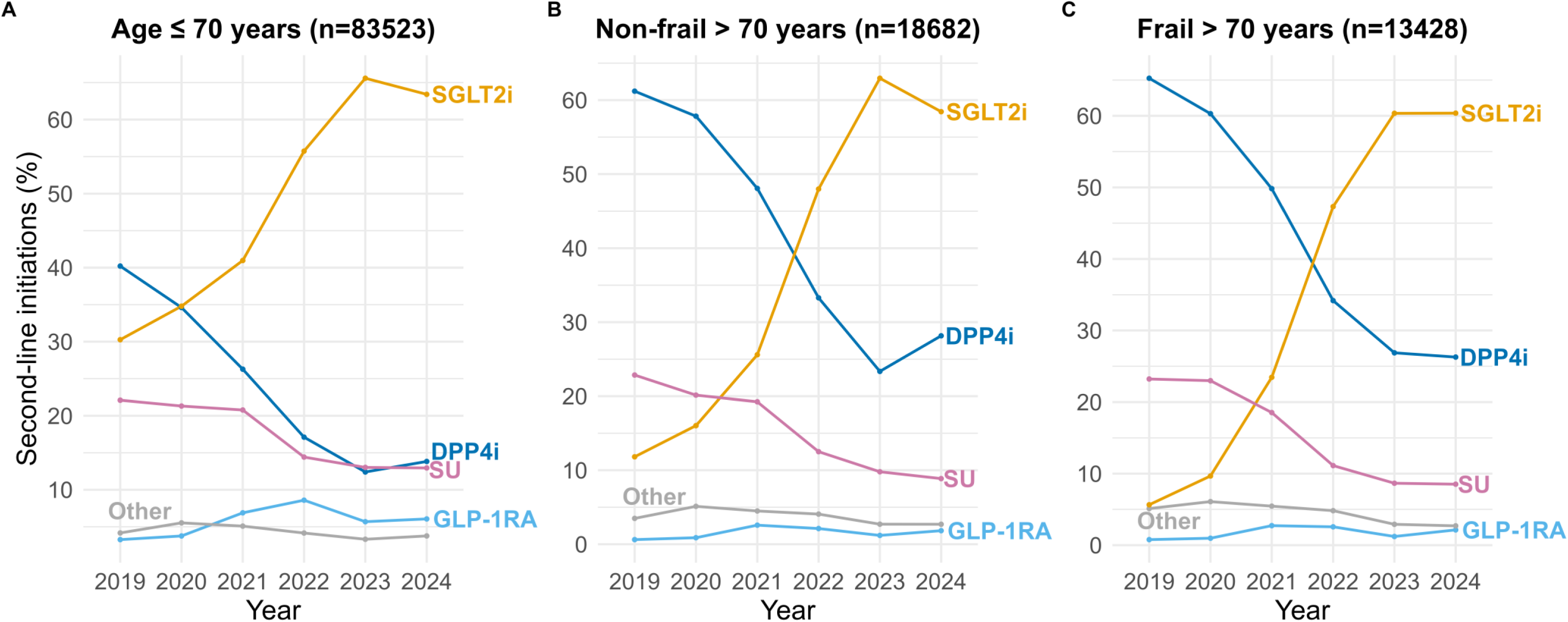
Time trends from 2019 to 2024* in new second-line drug therapy after metformin in patients (A) aged ≤70 years (n=84,589), (B) non-frail >70 years (n=18,933) and (C) frail >70 years (n=13,524). The prescriptions for each drug class each year are given as the percentage of total new drug prescriptions for second-line therapies that year. *2024 data include January to March.

During the study period, SGLT2i overtook DPP4i and sulfonylureas becoming the most common second-line therapy across all age and frailty groups, with DPP4i use declining overall from 44% of second-line initiations in 2019 to 16% in 2024 and SU use from 22% to 13%. We observed no increase in second-line GLP-1RA initiations from 2019 to 2024.

Prescribing trends were consistent across subgroups defined by sex, ethnicity, deprivation, among patients with or without pre-existing CVD or CKD and across frailty subgroups further stratified by moderate and severe frailty, with SGLT2i the most initiated therapy for all subgroups in 2024 (Figure S1-S6).

### Changes in prescribing were temporally associated with modest improvements in HbA_1c_ and weight response across age and frailty subgroups, without change in treatment discontinuation

Over 2019–2023, we found modest improvements in glycaemic response and weight change following second-line therapy initiation across all age and frailty subgroups (Figure 2A and 2B). In patients aged ≤70 years, mean 12-month HbA_1c_ response following initiation of second-line therapy improved from -11.3 mmol/mol (95% CI -11.6 to -11.0) in 2019 to -12.9 mmol/mol (95% CI -13.2 to -12.6) in 2023, an improvement of -1.6 mmol/mol (95% CI -2.0 to -1.2; p<0.001 for 2023 vs. 2019 difference; Figure 2A). Similar trends were also seen in frail and non-frail patients >70 years, although average response was greater in patients aged ≤70 years.

**Figure 2.**
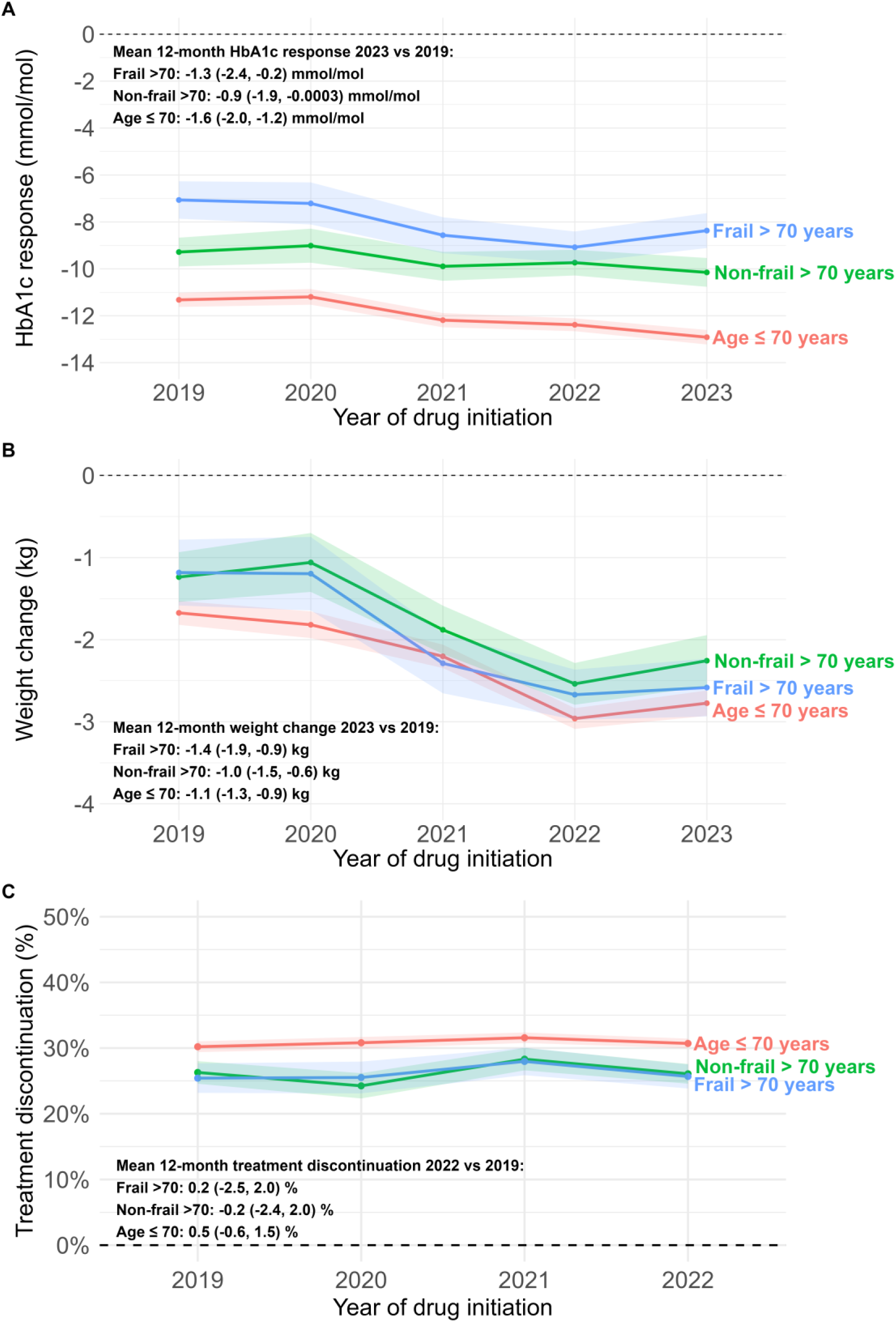
Adjusted mean 12-month response following second-line therapy initiation for (A) HbA1c (mmol/mol) and (B) weight (kg) 2019 to 2023 and (C) treatment discontinuation within 12-months of second-line therapy initiation 2019–2022, by age and frailty subgroups: aged ≤70 years (n=84,589), non-frail >70 years (n=18,933) and frail >70 years (n=13,524). Shading represents 95% confidence intervals.

Weight reductions over the same period followed a similar trend (Figure 2B). In patients aged ≤70 years, mean 12-month weight change following initiation of second-line therapy improved from -1.7 kg (95% CI -1.8 to -1.5) in 2019 to -2.8 kg (95% CI -2.9 to -2.6) in 2023, a difference of -1.1 kg (95% CI -1.3 to -0.9; p<0.001 for 2023 vs. 2019 difference). Similar trends were observed in frail and non-frail patients >70 years.

The risk of treatment discontinuation following initiation of second-line therapy remained stable (Figure 2C). In patients aged ≤70 years risk of discontinuation following second-line therapy initiation was 30.2% (95% CI 29.4 to 31.0) in 2019 compared to 30.7% in 2022 (95% CI 30.0 to 31.4), a difference in risk of 0.5% (95% CI -0.6 to 1.6; p=0.36 for 2022 vs 2019 difference). We observed no evidence of change in treatment discontinuation for frail and non-frail patients >70 years.

Time trends for glycaemic response, weight change and treatment discontinuation were consistent in sensitivity analyses using a 6-month instead of a 12-month window (Figure S7), and when further stratifying frailty status by moderate and severe frailty (Figure S8).

### Limited evidence of changes over time in incidence of short-term severe diabetes complications, heart failure, kidney failure, or DKA

In adults aged ≤70 years, the adjusted incidence rate of severe diabetes-related complications was stable over 2019–2022, 2019: 9.9 (95% CI 8.1 to 11.7; p<0.001); 2022: 8.6 (95% CI 6.9 to 10.3) per 1000 person-years; difference: -1.3, 95% CI -3.8 to 1.2; p=0.31; Figure 3A). Heart failure incidence was similarly stable (2019: 2.0 (95% CI 1.1 to 2.9), 2022: 2.7 (95% CI 1.8 to 3.7); difference: 0.7, 95% CI -0.5 to 2.0; p=0.20; Figure 3B), including in those with pre-existing heart failure or cardiovascular disease (Figure S9). Kidney failure incidence was also stable (2019: 2.1 (95% CI 1.1 to 3.0); 2022: 1.2 (95% CI 0.5 to 1.8); difference -0.9, -2.0 to 0.3; p=0.13; Figure 3C). Although adjusted incidence rates were higher for all outcomes, stable trends by calendar year were consistently observed in frail and non-frail patients >70 years.

**Figure 3.**
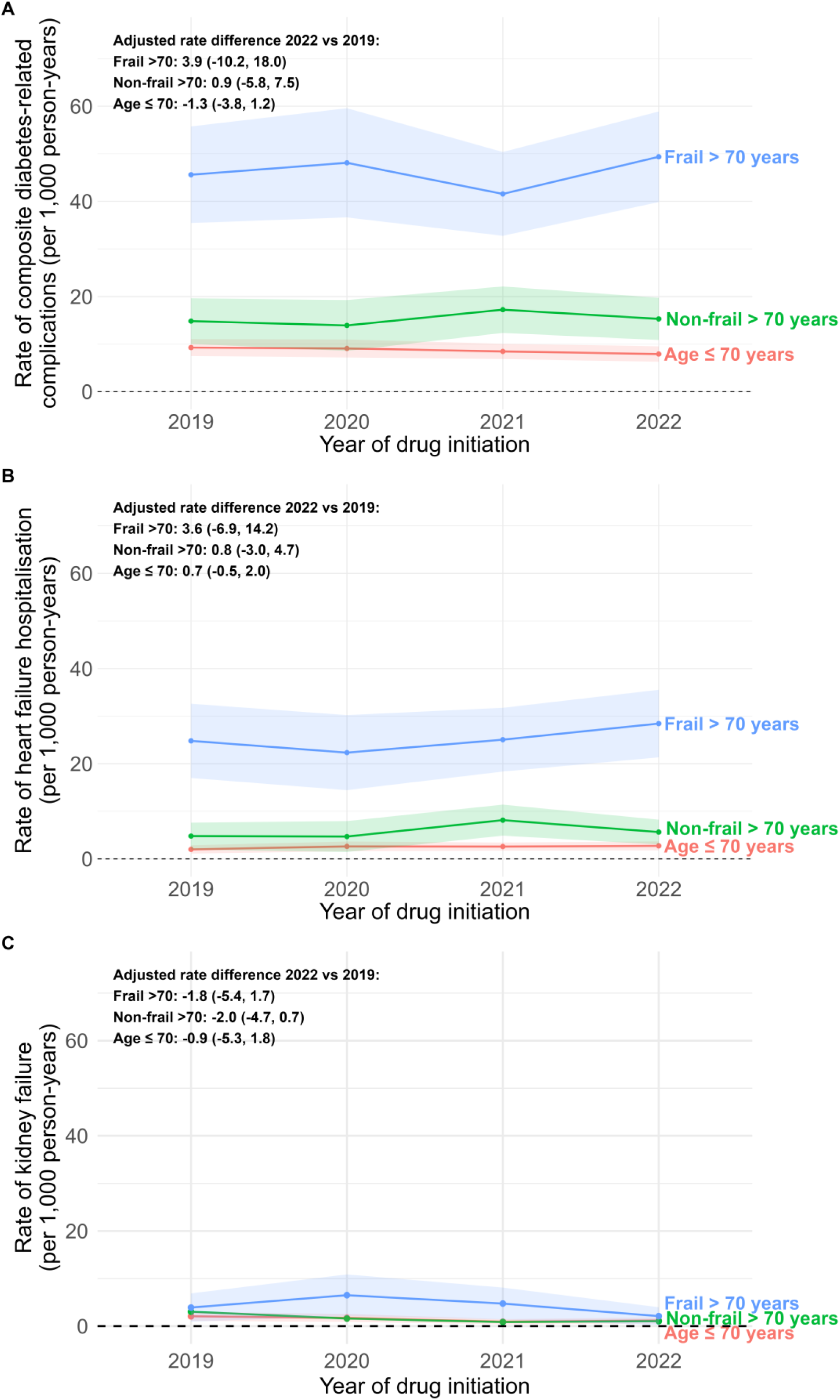
Adjusted incidence rates of (A) a severe diabetes-related complication (sudden death; death due to hyperglycaemia or hypoglycaemia; fatal or non-fatal myocardial infarction, angina, heart failure, stroke, or kidney failure; death from peripheral vascular disease; amputation; blindness; and severe retinopathy [vitreous haemorrhage or retinal photocoagulation]), (B) heart failure, and (C) kidney failure following initiation of second-line therapy, stratified by age and frailty: ≤70 years (n = 84,589), non-frail >70 years (n = 18,933), and frail >70 years (n = 13,524), 2019 to 2022. Shaded areas represent 95% confidence intervals. Underlying crude incidence rates for all complication outcomes are reported in Supplementary Table 3.

Crude incidence of DKA remained rare across all subgroups (<50 events total across subgroups from 2019 to 2022) no evidence of meaningful change by calendar year (Supplementary Table 3).

### Impact of UK NICE 2022 guideline changes on SGLT2i initiation

Overall, monthly SGLT2i initiations were already increasing before the 2022 NICE guidelines by 0.55% per month (95% CI 0.35 to 0.74; p<0.001) (Figure 4). In the month following the publication of the guidelines, there was an immediate 7.5% increase in monthly initiations (95% CI 3.1 to 11.9; p<0.001). In the months after that, the increase in monthly initiations continued at 0.76% per month, an additional 0.21% per month compared to the pre-guideline trend (95% CI -0.21 to 0.62; p=0.33). We observed similar trends in both frail and non-frail adults >70 years, though the immediate increase in SGLT2i initiations in the month following the publication of the NICE 2022 guidelines was greater in both non-frail >70 years (11.5% [95% CI 5.5 to 17.4; p<0.001]) and frail >70 years (14.5% [95% CI 7.7 to 21.5; p<0.001]) adults (Figure S10). Shading represents 95% confidence intervals.

**Figure 4.**
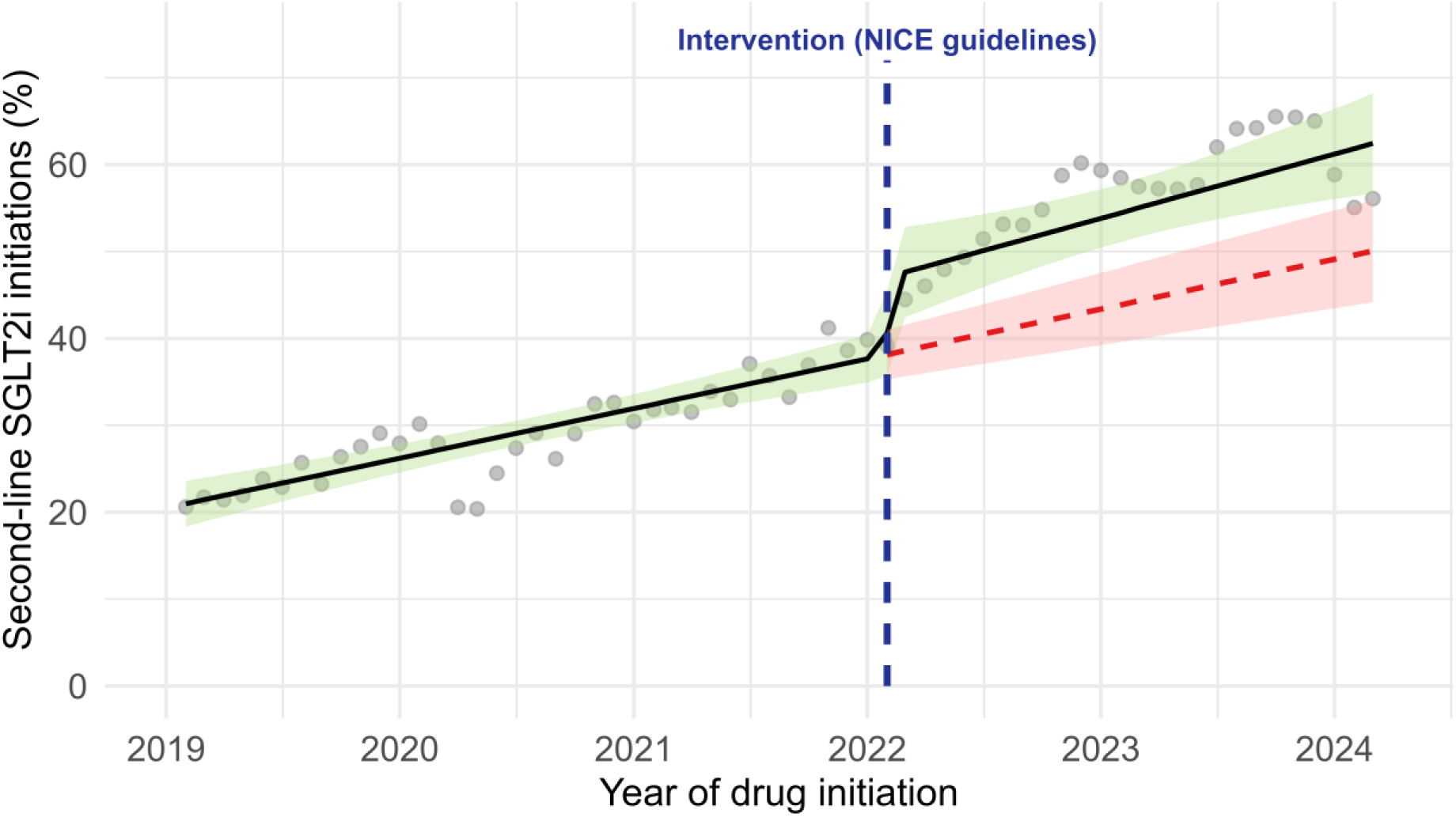
Interrupted time series analysis of monthly SGLT2 inhibitor initiations, 2019-2024. The solid black line shows the observed trend, the dashed red line shows the counterfactual trend, and the vertical dashed blue line shows the intervention (publication of the NICE guidelines).

## DISCUSSION

This population-based cohort study reveals a major shift in initiations of second-line glucose-lowering therapies in the UK between 2019 and 2024, with SGLT2i becoming the most common second-line therapy across all age and frailty subgroups. SGLT2i have largely replaced DPP4i and sulfonylureas. Over the same time period we observed modest population-level improvements in HbA_1c_ response and weight loss after treatment initiation, but no evidence of changes in short-term treatment discontinuation or rates of severe diabetes related complications, heart failure, kidney failure, or DKA within 12-months of treatment initiation. Importantly, these trends in short-term outcomes were consistent in frail older adults despite the proportion of SGLT2i initiations increasing 10-fold in this group.

SGLT2i have become the most commonly initiated second-line therapy in the UK. This represents a clear shift from earlier UK studies, which found DPP4i to be the most common second-line therapy overall and in people with cardiorenal comorbidity, despite an absence of cardiorenal benefit and comparable cost to SGLT2i.^6, 15 16^ The immediate increase in SGLT2i initiations following the 2022 NICE update suggests that national guidance can rapidly influence clinical practice. Encouragingly, this increase was seen across all other population subgroups evaluated, including by ethnicity, deprivation, and amongst people with CVD and CKD. In contrast to the marked change in SGLT2i initiations, we did not observe a corresponding increase in second-line GLP-1RA use, despite trial-based evidence of cardiovascular and renal benefit.^30^ This likely reflects their later placement in UK guidance, where GLP-1RA are primarily initiated as third line therapies and limited to individuals with a BMI ≥35 kg/m².^4^

The modest population-level improvements in HbA_1c_ and weight are consistent with the replacement of DPP4i prescribing for SGLT2i. Clinical trials have consistently shown greater glycaemic and weight benefits with SGLT2i compared with DPP4i and the effects observed in our study (mean HbA_1c_ reduction of -1.6 mmol/mol and mean weight loss of -1.1 kg) align with these findings.^31, 32 33 34 35 36^ Although modest at the individual level, a - 1.6mmol/mol greater average HbA_1c_ reduction has meaningful population-level implications. Given that the average rate of glycaemic progression in type 2 diabetes is approximately 1 mmol/mol per year, a 1.6 mmol/mol benefit translates to an average 1.6 additional years of stable glycaemic control before treatment intensification may be required.^37^ In the future, implementation of precision medicine approaches, such as a recently developed model to predict optimal glucose-lowering therapy considering 5 major drug classes after metformin,^38^ could provide an alternative to current prescribing practice by allowing targeting of therapy to optimise glycaemic response for an individual patient based on their routine clinical features.

Despite the marked increase in SGLT2i prescribing, we did not find evidence of meaningful population-level changes in short-term severe diabetes complications including incidence of hospitalisation for heart failure or end stage kidney failure, despite the known benefits of SGLT2i for these outcomes.^39^ Several factors may explain this. First, as our focus was on evaluating temporal trends, complication outcomes were limited by a 12-month follow-up window following treatment second-line initiation. Although cardiovascular and renal benefits of SGLT2i have been observed in trials within the first year,^40^ participants have been predominantly those with pre-existing cardiovascular and kidney disease in contrast to our lower risk population-based cohort. Supporting this, a recent meta-analysis found the absolute risk reduction with SGLT2i for hospitalisation for heart failure was substantially greater in those with established cardiovascular disease than those without.^41^ The unavailability of hospital admission data beyond 2023 also meant we lacked data on complication outcomes during the period of highest SGLT2i uptake. Second, because our outcome analyses included all second-line therapies, not just SGLT2i, even in the most recent years any overall treatment effects may have been attenuated by the 40% of patients who initiated alternative drug classes without cardiovascular and renal benefits. Third, the study period overlapped with the COVID-19 pandemic which likely affected hospital admissions. Our analysis therefore highlights the need for focused studies evaluating longer-term real-world outcomes with SGLT2i as their use across the wider population with type 2 diabetes increases.

DKA is a recognised adverse event with SGLT2i, prompting safety warnings from the European Medicines Agency.^42^ In our study, we found no increase in DKA incidence following initiation of second-line therapy, with events remaining rare even in frail older adults. However, recent UK causal analysis suggests that although absolute rates remain low, the relative risk of DKA is higher with SGLT2i in adults aged ≥70 years compared with DPP4i, but not in those <70 years,^24^ highlighting the importance of careful prescribing and monitoring in older adults.

Our study has several strengths. We used large, population-based data from CPRD, which is broadly representative of the UK population by age, sex, and ethnicity.^43^ We applied a validated electronic frailty index, allowing robust subgroup analyses in frail populations who are often excluded from clinical trials.^19^ However, as an observational study, we cannot infer causality between prescribing changes, outcomes, and guideline updates. In addition, our adjusted analysis assumes that differences across calendar years reflect temporal effects alone, but unmeasured confounders including changes in healthcare delivery post-COVID may not have been fully accounted for.

In summary, from 2019 to 2024, there has been a substantial shift in second-line therapy for type 2 diabetes in the UK, with SGLT2i becoming the most commonly initiated treatment, and the greatest increase in SGLT2i use in older adults with frailty. Over the same period, short-term outcomes remained stable, with modest benefits for HbA_1c_ and weight and no meaningful change in treatment discontinuation or major complications including heart failure, kidney failure, or DKA. These findings support the careful use of SGLT2i across the wider population with type 2 diabetes including frail older adults.

## Data Availability

Access to CPRD data is subject to protocol approval via the CPRD research data governance process (https://cprd.com/data-access). Code for initial cohort preparation is available at: https://github.com/Exeter-Diabetes/CPRD-Cohort-scripts/tree/main/03-Treatment-response-(MASTERMIND) and analysis code is available at: https://github.com/Exeter-Diabetes/CPRD-Martha-prescribing-trends.

https://github.com/Exeter-Diabetes/CPRD-Martha-prescribing-trends

## Contributors

The study concept and design were conceived and developed by MMD, JMD, BMS, TJM and KGY. MMD, JMD, KGY, PC, LMG, TTJ, and BMS had access to all the raw datasets used for the study. APM reviewed the clinical codes. MMD undertook the analysis, with support from KGY, TJM, BMS and JMD. All authors provided support for the analysis and interpretation of results, critically revised the manuscript, and approved the final manuscript. MMD and JMD attests that all listed authors meet authorship criteria and that no others meeting the criteria have been omitted. All authors had final responsibility for the decision to submit for publication.

## Transparency

The lead author (JMD) is the guarantor of this manuscript and affirms that the manuscript is an honest, accurate, and transparent account of the study being reported; that no important aspects of the study have been omitted; and that any discrepancies from the study as originally planned have been explained. The study protocol was not pre-registered.

## Ethical approval

This study was approved by the CPRD independent scientific advisory committee (eRAP 24_004747). CPRD also has ethical approval from the Health Research Authority to support research using anonymised patient data (research ethics committee reference 21/EM/0265). Individual patient consent was not required as all data were deidentified.

## Data availability

Access to CPRD data is subject to protocol approval via CPRD’s research data governance process (https://cprd.com/data-access). Code for initial cohort preparation is available at: https://github.com/Exeter-Diabetes/CPRD-Cohort-scripts/tree/main/03-Treatment-response-(MASTERMIND) and analysis code is available at: https://github.com/Exeter-Diabetes/CPRD-Martha-prescribing-trends.

## Provenance and peer review

Not commissioned; externally peer reviewed

