## Supplementary Tables 1 for "Trends in second-line glucose-lowering therapy initiations and treatment outcomes across age and frailty groups in people with type 2 diabetes: UK population-based study, 2019-2024"

**Supplementary Table 1.** Baseline characteristics of the study cohort at second-line treatment initiation by calendar year (2019-2024).

|  | **2019** | **2020** | **2021** | **2022** | **2023** | **2024** |
| --- | --- | --- | --- | --- | --- | --- |
| Second line initiations | 19,354 | 16,391 | 20,918 | 24,632 | 29,569 | 6182 |
| Age at therapy initiation (years) | 61.4 (13.1) | 60.7 (13.2) | 61.5 (13.2) | 62.0 (13.2) | 62.1 (13.2) | 62.1 (13.2) |
| Sex (% Male) | 11,356 (58.7) | 9615 (58.7) | 12,163 (58.1) | 14,549 (59.1) | 17,635 (59.6) | 3763 (60.9) |
| Duration of diabetes (years) | 6.0 (4.1) | 5.9 (4.8) | 6.2 (5.0) | 6.4 (5.0) | 6.4 (5.3) | 6.5 (5.4) |
| **Ethnicity (%)** | | | | | | |
| White | 15,520 (80.2) | 13,085 (79.8) | 16,873 (80.7) | 19,881 (80.7) | 23,480 (79.4) | 4840 (78.3) |
| South Asian | 2338 (12.1) | 1886 (11.5) | 2296 (11.0) | 2727 (11.1) | 3577 (12.1) | 782 (12.6) |
| Black | 759 (3.9) | 763 (4.7) | 992 (4.7) | 1076 (4.4) | 1338 (4.5) | 313 (5.1) |
| Mixed | 189 (1.0) | 179 (1.1) | 208 (1.0) | 219 (0.9) | 299 (1.0) | 54 (0.9) |
| Other | 278 (1.4) | 257 (1.6) | 307 (1.4) | 497 (2.0) | 516 (1.7) | 104 (1.7) |
| Missing | 270 (1.4) | 221 (1.3) | 242 (1.2) | 232 (0.9) | 359 (1.2) | 89 (1.4) |
| **IMD quintile (%)** | | | | | | |
| 1 (least deprived) | 2565 (13.3) | 2074 (12.7) | 2848 (13.6) | 3330 (13.5) | 3916 (13.2) | 826 (13.4) |
| 2 | 2924 (15.1) | 2436 (14.9) | 3037 (14.5) | 3644 (14.8) | 4243 (14.3) | 893 (14.4) |
| 3 | 3002 (15.5) | 2425 (14.8) | 3149 (15.1) | 3808 (15.5) | 4539 (15.4) | 952 (15.4) |
| 4 | 3465 (17.9) | 3014 (18.4) | 3805 (18.2) | 4358 (17.7) | 5113 (17.3) | 1026 (16.6) |
| 5 (most deprived) | 3828 (19.8) | 3352 (20.5) | 4269 (20.4) | 4808 (19.5) | 5589 (18.9) | 1155 (18.7) |
| Missing | 3570 (18.4) | 3090 (18.7) | 3810 (18.2) | 4684 (9.0) | 6169 (20.9) | 1330 (21.5) |
| **Clinical features** | | | | | | |
| BMI (kg/m²) | 32.8 (7.2) | 33.1 (7.3) | 33.3 (7.5) | 33.3 (7.6) | 33.1 (7.5) | 33.0 (7.4) |
| Weight (kg) | 93.6 (22.5) | 94.9 (22.7) | 95.2 (23.1) | 95.0 (23.6) | 94.7 (23.1) | 94.5 (23.2) |
| HbA1c (mmol/mol) | 72.3 (19.1) | 76.0 (20.8) | 75.2 (20.7) | 72.1 (19.9) | 69.7 (19.8) | 70.0 (20.0) |
| **Comorbidities (%)** |  |  |  |  |  |  |
| CVD^a^ | 4531 (23.4) | 3901 (23.8) | 5271 (25.5) | 6295 (25.6) | 7368 (24.9) | 1487 (24.1) |
| CKD stage 3-4 | 2226 (11.5) | 1788 (10.9) | 2400 (11.5) | 2898 (11.8) | 3222 (10.9) | 620 (10.0) |
| CKD stage 5 | 70 (0.4) | 56 (0.3) | 70 (0.3) | 85 (0.3) | 92 (0.3) | 17 (0.3) |
| Heart failure | 1410 (7.3) | 1306 (8.0) | 2044 (9.8) | 2760 (11.2) | 2951 (10.0) | 566 (9.2) |
| Hyperglycaemia | 37 (0.2) | 52 (0.3) | 53 (0.3) | 55 (0.2) | 25 (0.1) | 2 (0.0) |
| Hypoglycaemia | 13 (0.1) | 13 (0.1) | 20 (0.1) | 21 (0.1) | 10 (0.0) | 2 (0.0) |
| Lower limb amputation | 42 (0.2) | 46 (0.3) | 73 (0.3) | 61 (0.2) | 65 (0.2) | 13 (0.2) |
| Severe retinopathy^b^ | 57 (0.3) | 69 (0.4) | 70 (0.3) | 94 (0.4) | 107 (0.4) | 16 (0.3) |
| **Drug class (%)** |  |  |  |  |  |  |
| DPP4i | 8975 (46.4) | 6678 (40.7) | 6786 (32.4) | 5384 (21.9) | 4727 (16.0) | 1091 (17.6) |
| SGLT2i | 4756 (24.6) | 4814 (29.4) | 7640 (36.5) | 13,162 (53.4) | 19,074 (64.5) | 3847 (62.2) |
| GLP-1RA | 495 (2.6) | 498 (3.0) | 1197 (5.7) | 1672 (6.8) | 1299 (4.4) | 303 (4.9) |
| SU | 4324 (22.3) | 3495 (21.3) | 4242 (20.3) | 3375 (13.7) | 3534 (12.0) | 727 (11.8) |
| Other | 804 (4.2) | 906 (5.5) | 1053 (5.0) | 4.2 (4) | 935 (3.2) | 214 (3.5) |

Values for continuous variables are given as mean +-SD and binary variables as n (%)

^a^CVD: myocardial infarction, stroke, revascularisation, ischaemic heart disease, angina, peripheral arterial disease, transient ischaemic attack. ^b^Severe retinopathy: vitreous haemorrhage, retinal photocoagulation

**Supplementary Table 2.** Baseline characteristics of the study cohort at second-line treatment initiation with age and frailty subgroups further stratified into moderate and severe frailty.

| **Variable** | **Age ≤ 70 years** | **Non-frail > 70 years** | **Moderate > 70 years** | **Severe > 70 years** |
| --- | --- | --- | --- | --- |
| N (%) | 84,589 (72.3) | 18,933 (16.2) | 8961 (7.6) | 4563 (3.9) |
| Age at therapy (years) | 55.5 (9.6) | 76.2 (4.7) | 78.9 (5.7) | 81.5 (6.2) |
| Sex (% Male) | 50,489 (59.7) | 11,625 (61.4) | 4889 (54.6) | 2018 (44.4) |
| Duration of diabetes (years) | 5.1 (4.3) | 8.4 (5.4) | 9.8 (5.8) | 11.0 (6.0) |
| **Ethnicity (%)** |  |  |  |  |
| White | 64,398 (76.1) | 17,063 (90.1) | 8073 (90.1) | 4145 (90.8) |
| South Asian | 11,959 (14.1) | 851 (4.5) | 528 (5.9) | 268 (5.9) |
| Black | 4578 (5.4) | 391 (2.1) | 185 (2.1) | 87 (1.9) |
| Mixed | 992 (1.2) | 103 (0.5) | 45(0.7) | 8 (0.5) |
| Other | 1596 (1.9) | 274 (1.4) | 66 (0.5) | 23 (0.2) |
| Missing | 1066 (1.3) | 251 (1.3) | 64 (0.7) | 32 (0.7) |
| **IMD quintile (%)** |  |  |  |  |
| 1 (least deprived) | 9810 (11.6) | 3584 (18.9) | 1498 (16.7) | 667 (14.6) |
| 2 | 11,34 (13.4) | 3531 (18.6) | 1593 (17.8) | 719 (15.8) |
| 3 | 12,630 (14.9) | 3083 (16.3) | 1459 (16.3) | 703 (15.4) |
| 4 | 15,830 (18.7) | 2727 (14.4) | 1458 (16.3) | 765 (16.8) |
| 5 (most deprived) | 18,463 (21.8) | 2338 (12.3) | 1352 (15.1) | 848 (18.6) |
| Missing | 16,521 (19.6) | 3670 (19.5) | 1606 (17.8) | 861 (18.8) |
| **Clinical features** |  |  |  |  |
| BMI (kg/m²) | 34.2 (7.7) | 30.3 (5.9) | 30.6 (6.3) | 30.5 (6.6) |
| Weight (kg) | 98.5 (23.5) | 85.4 (17.8) | 84.6 (18.8) | 82.0 (19.0) |
| HbA1c (mmol/mol) | 74.3 (20.3) | 68.7 (18.5) | 67.0 (19.6) | 65.3 (20.2) |
| **Comorbidities (%)** |  |  |  |  |
| CVD^a^ | 15,091 (17.8) | 5397 (28.5) | 5006 (55.9) | 3359 (73.6) |
| CKD stage 3-4 | 2709 (3.2) | 4191 (22.1) | 3638 (40.6) | 2616 (57.3) |
| CKD stage 5 | 258 (0.3) | 49 (0.3) | 52 (0.6) | 31 (0.7) |
| Heart failure | 4485 (5.3) | 1607 (8.6) | 2518 (28.1) | 2345 (52.0) |
| Hyperglycaemia | 171 (0.2) | 22 (0.1) | 18 (0.2) | 12 (0.2) |
| Hypoglycaemia | 52 (0.1) | 12 (0.1) | 10 (0.1) | 5 (0.1) |
| Lower limb amputation | 179 (0.2) | 38 (0.2) | 33 (0.3) | 47 (1.0) |
| Severe retinopathy^b^ | 192 (0.2) | 109 (0.6) | 69 (0.8) | 43 (1.0) |
| **eFI category (%)** |  |  |  |  |
| Fit | 45,328 (53.6) | 5554 (29.3) |  |  |
| Mild | 30,701 (36.3) | 13,379 (70.6) |  |  |
| Moderate | 7227 (8.5) |  | 8961 (100) |  |
| Severe | 1333 (1.6) |  |  | 4563 (100) |

Values for continuous variables are given as mean +-SD and binary variables as n (%)

^a^CVD: myocardial infarction, stroke, revascularisation, ischaemic heart disease, angina, peripheral arterial disease, transient ischaemic attack

^b^Severe retinopathy: vitreous haemorrhage, retinal photocoagulation

**Supplementary Figure 1.** Trends in second-line initiations by sex (2019-2024)

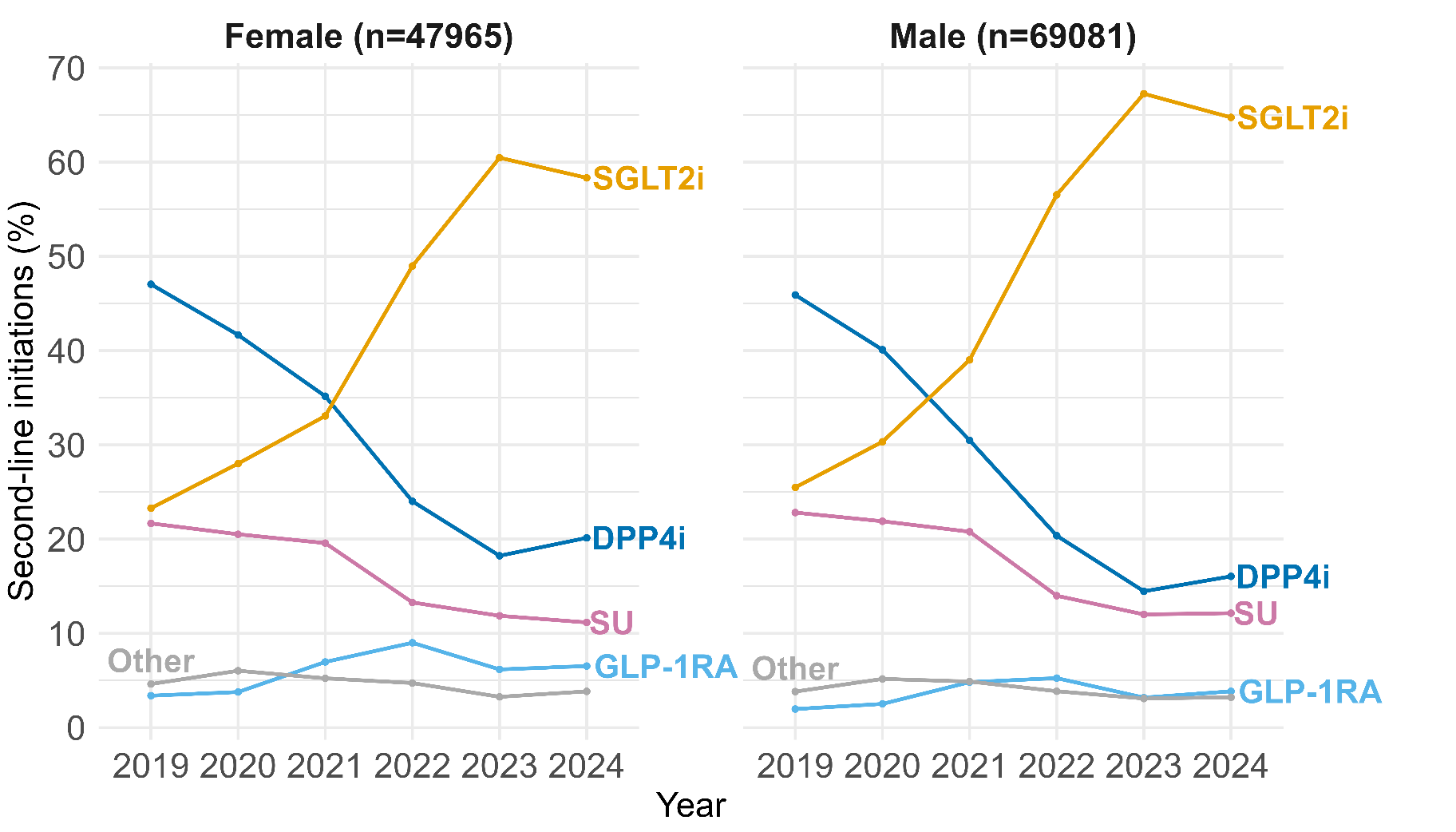

**Supplementary Figure 2.** Trends in second-line initiations by ethnicity (2019-2024)

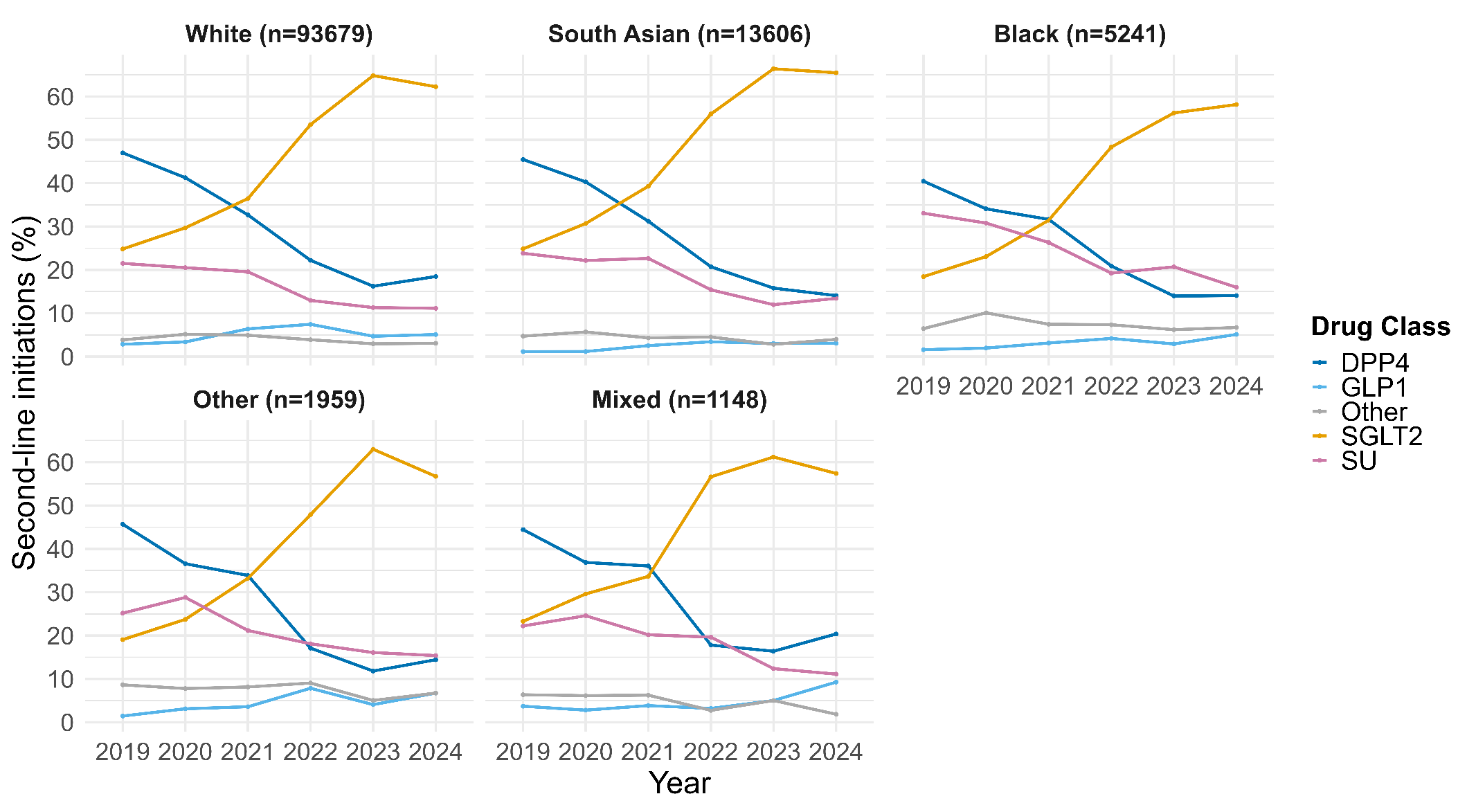

**Supplementary Figure 3**. Trends in second-line initiations by deprivation (IMD quintiles) (2019–2024)

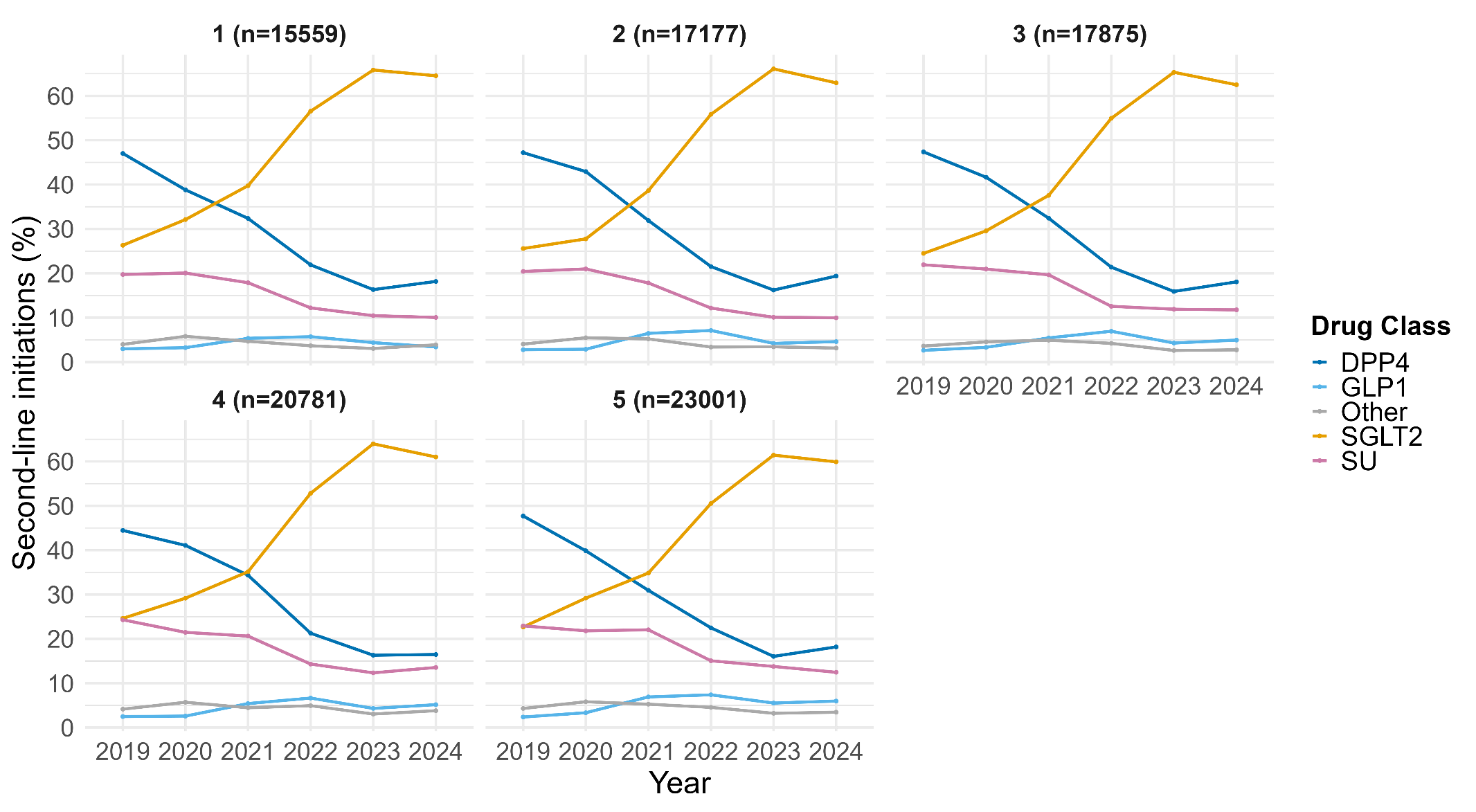

**Supplementary Figure 4.** Trends in second-line initiations by baseline cardiovascular disease status (2019-2024).

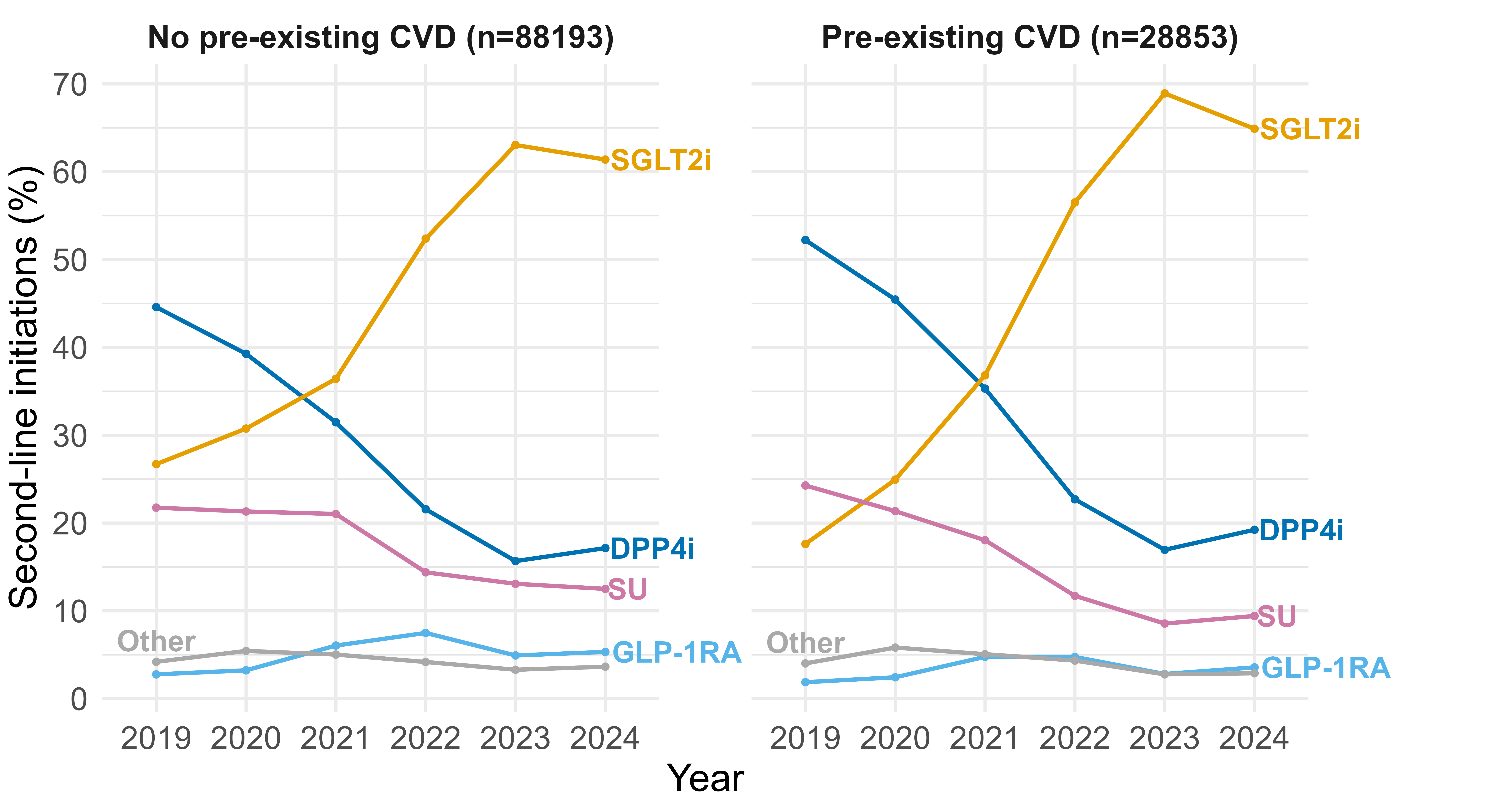

**Supplementary Figure 5.** Trends in second-line initiations by baseline chronic kidney disease status (2019-2024).

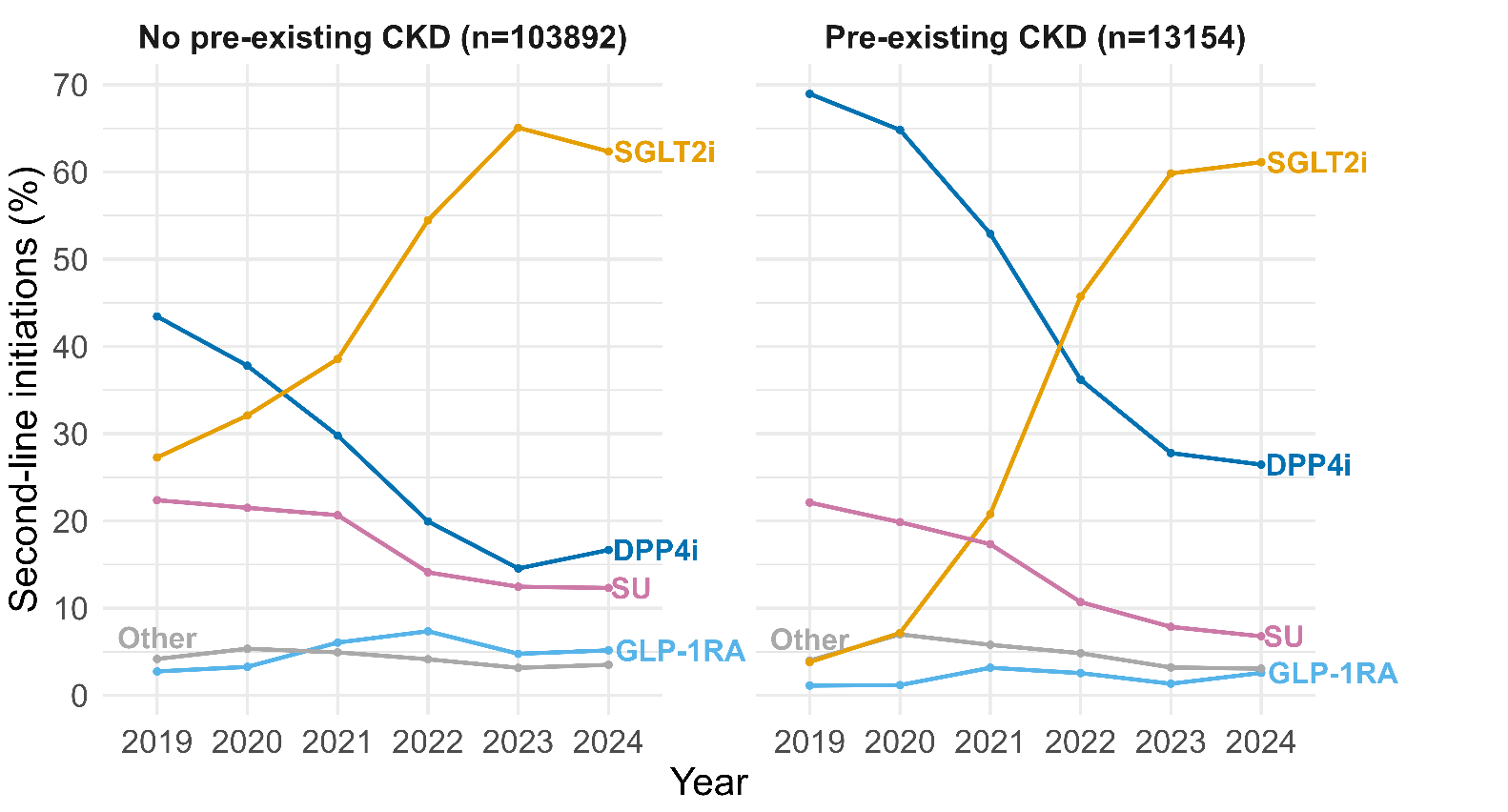

**Supplementary Figure 6.** Trends in second-line initiations by age and frailty subgroup (2019-2024), with frailty further categorised into moderate and severe**.**

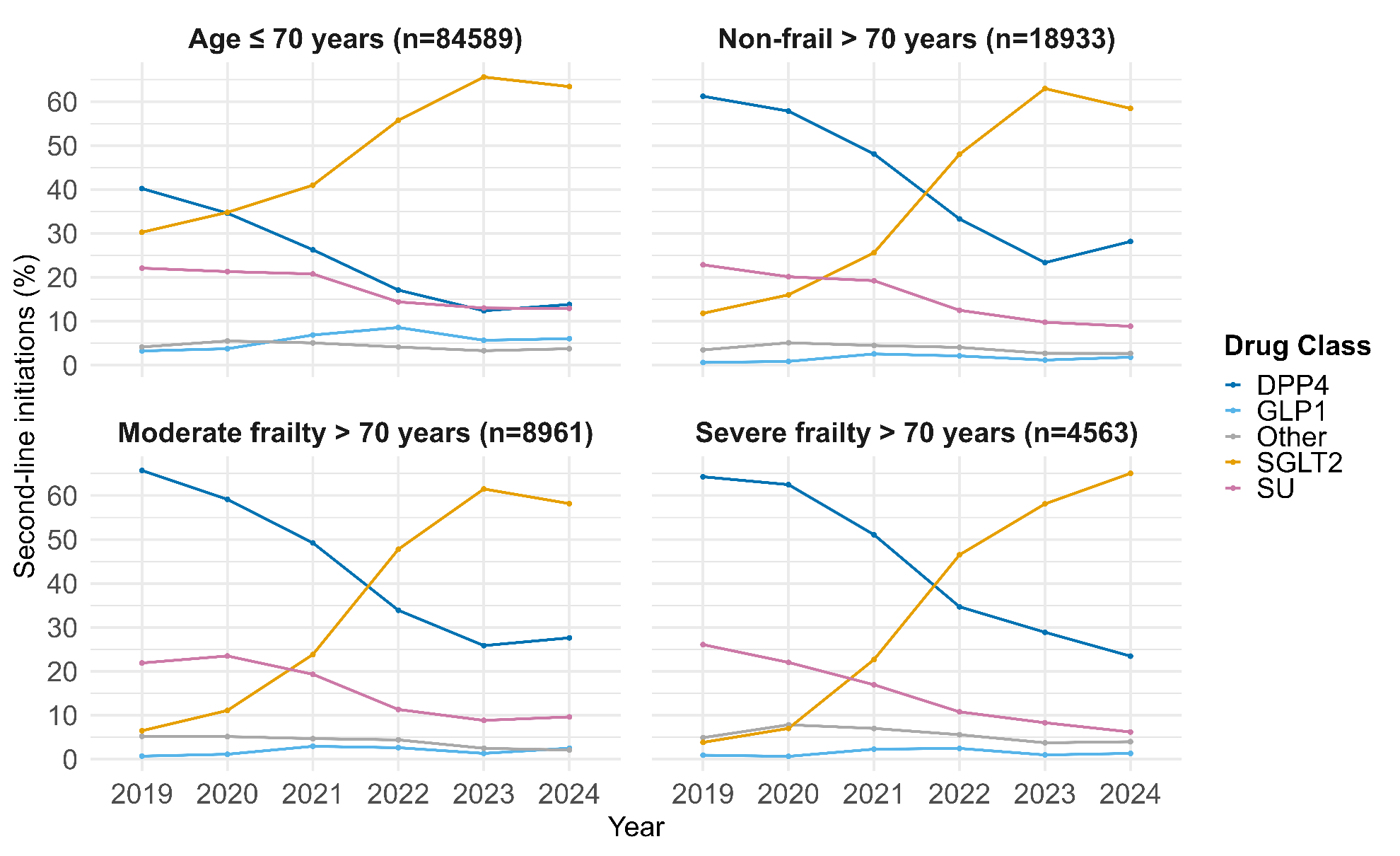

**Supplementary Figure 7.** 6-month HbA1c response, weight change (2019-2023) and treatment discontinuation (2019-2022) following second-line therapy initiation by age and frailty subgroups.

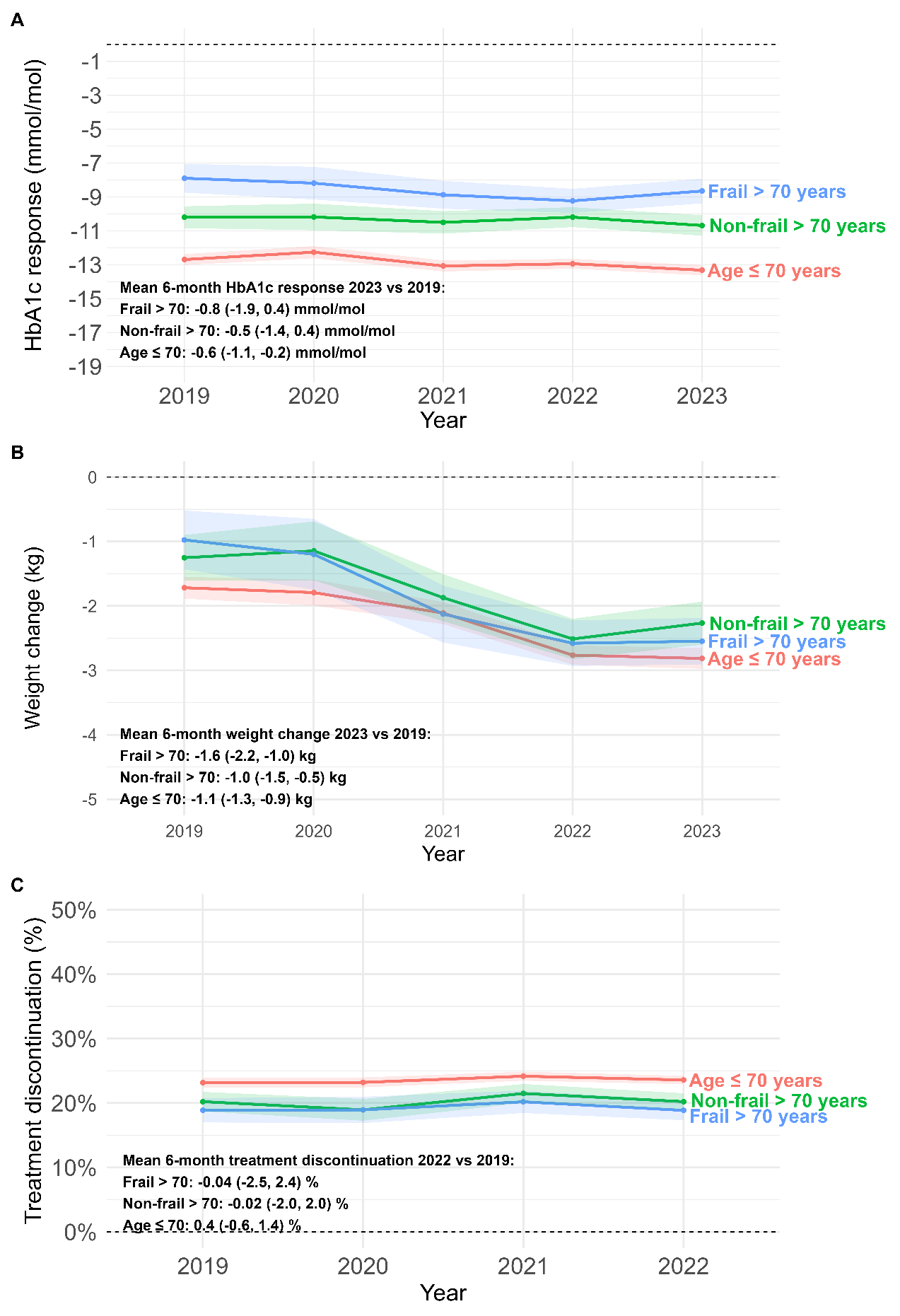

**Supplementary Figure 8.** 12-month HbA1c response, weight change (2019-2023) and treatment discontinuation (2019-2022) following second-line therapy initiation by age and frailty subgroups, with frailty further categorised into moderate and severe.

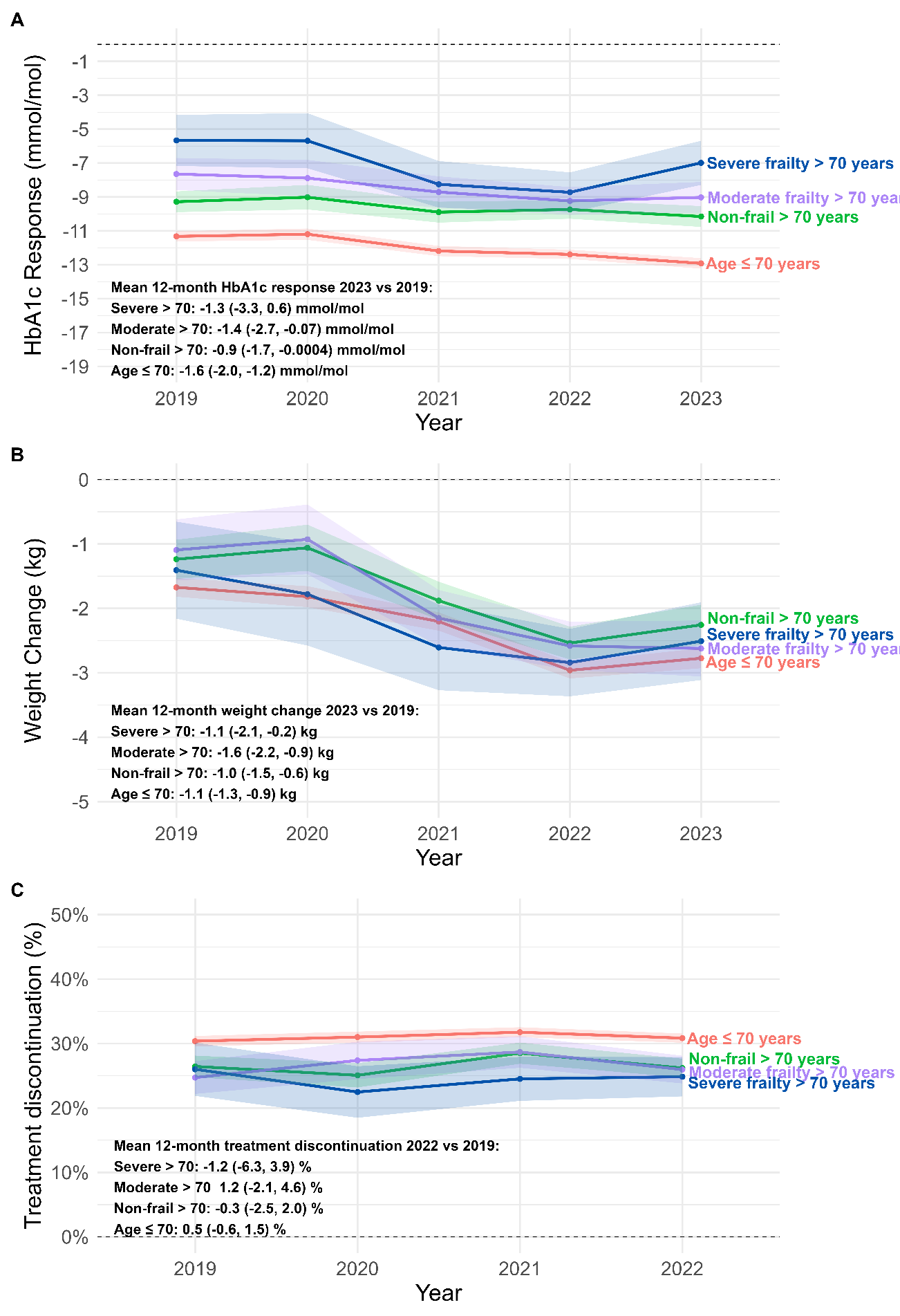

**Supplementary Table 3.** Crude incidence rates of complications following second-line therapy initiation including a severe diabetes-related complications, heart failure, kidney failure and DKA, per 1000 person-years by calendar year (2019–2022) and frailty category a) age ≤ 70 (n= 84,589) b) non-frail > 70 (n=18,933) c) frail > 70 (n=13,542).

**a) Age ≤ 70 years**

| **Outcome** | **Year** | **n** | **No. of outcome events** | **Crude incidence rate /1000 person-years** |
| --- | --- | --- | --- | --- |
| **Severe diabetes-related complications^a^** | 2019 | 11367 | 111 | 12.9 |
|  | 2020 | 9892 | 99 | 13.3 |
|  | 2021 | 12327 | 117 | 13.0 |
|  | 2022 | 14153 | 99 | 13.0 |
| **Heart failure^b^** | 2019 | 11367 | 20 | 2.3 |
|  | 2020 | 9892 | 25 | 3.4 |
|  | 2021 | 12327 | 35 | 3.9 |
|  | 2022 | 14153 | 33 | 4.3 |
| **Kidney failure^c^** | 2019 | 11367 | 20 | 2.3 |
|  | 2020 | 9892 | 18 | 2.4 |
|  | 2021 | 12327 | 12 | 1.3 |
|  | 2022 | 14153 | 14 | 1.8 |
| **DKA** | 2019 | 11367 | 5 | 0.6 |
|  | 2020 | 9892 | 8 | 1.1 |
|  | 2021 | 12327 | 11 | 1.2 |
|  | 2022 | 14153 | 5 | 0.7 |

**b) Non-frail > 70 years**

| **Outcome** | **Year** | **n** | **No. of outcome events** | **Crude incidence rate /1000 person-years** |
| --- | --- | --- | --- | --- |
| **Severe diabetes-related complications^a^** | 2019 | 2613 | 38 | 19.0 |
|  | 2020 | 1904 | 28 | 19.2 |
|  | 2021 | 2728 | 51 | 25.4 |
|  | 2022 | 3279 | 48 | 27.0 |
| **Heart failure^b^** | 2019 | 2613 | 11 | 5.5 |
|  | 2020 | 1904 | 8 | 5.4 |
|  | 2021 | 2728 | 24 | 11.9 |
|  | 2022 | 3279 | 17 | 9.5 |
| **Kidney failure^c^** | 2019 | 2613 | 6 | 3.0 |
|  | 2020 | 1904 | 3 | 2.0 |
|  | 2021 | 2728 | 2 | 1.0 |
|  | 2022 | 3279 | 4 | 2.2 |
| **DKA** | 2019 | 2613 | n < 5 | 0.0 |
|  | 2020 | 1904 | n < 5 | 0.7 |
|  | 2021 | 2728 | n < 5 | 1.0 |
|  | 2022 | 3279 | n < 5 | 1.1 |

**c) Frail > 70 years**

| **Outcome** | **Year** | **n** | **No. of outcome events** | **Crude incidence rate /1000 person-years** |
| --- | --- | --- | --- | --- |
| **Severe diabetes-related complications^a^** | 2019 | 1715 | 80 | 64.2 |
|  | 2020 | 1436 | 69 | 68.4 |
|  | 2021 | 1972 | 87 | 62.1 |
|  | 2022 | 2416 | 106 | 82.8 |
| **Heart failure^b^** | 2019 | 1715 | 39 | 30.9 |
|  | 2020 | 1436 | 29 | 28.3 |
|  | 2021 | 1972 | 54 | 38.1 |
|  | 2022 | 2416 | 61 | 47.2 |
| **Kidney failure^c^** | 2019 | 1715 | 7 | 5.5 |
|  | 2020 | 1436 | 9 | 8.7 |
|  | 2021 | 1972 | 8 | 5.6 |
|  | 2022 | 2416 | 5 | 3.8 |
| **DKA** | 2019 | 1715 | n < 5 | 0.0 |
|  | 2020 | 1436 | n < 5 | 0.0 |
|  | 2021 | 1972 | 6 | 4.2 |
|  | 2022 | 2416 | n < 5 | 0.8 |

^a^ Severe diabetes-related complication: sudden death, death from hyperglycaemia or hypoglycaemia, fatal or non-fatal myocardial infarction, angina, fatal or non-fatal heart failure, fatal or non-fatal stroke, fatal or non-fatal kidney failure, death from peripheral vascular disease, amputation, blindness and severe retinopathy (vitreous haemorrhage, retinal photocoagulation)

^b^Heart failure: Fatal or non-fatal heart failure

^c^Kidney failure: Fatal or non-fatal kidney failure

**Supplementary Figure 9.** Rate (per 1000 person-years) following second-line therapy initiation of heart failure hospitalisations in patients with pre-existing heart failure or cardiovascular disease across age and frailty subgroups.

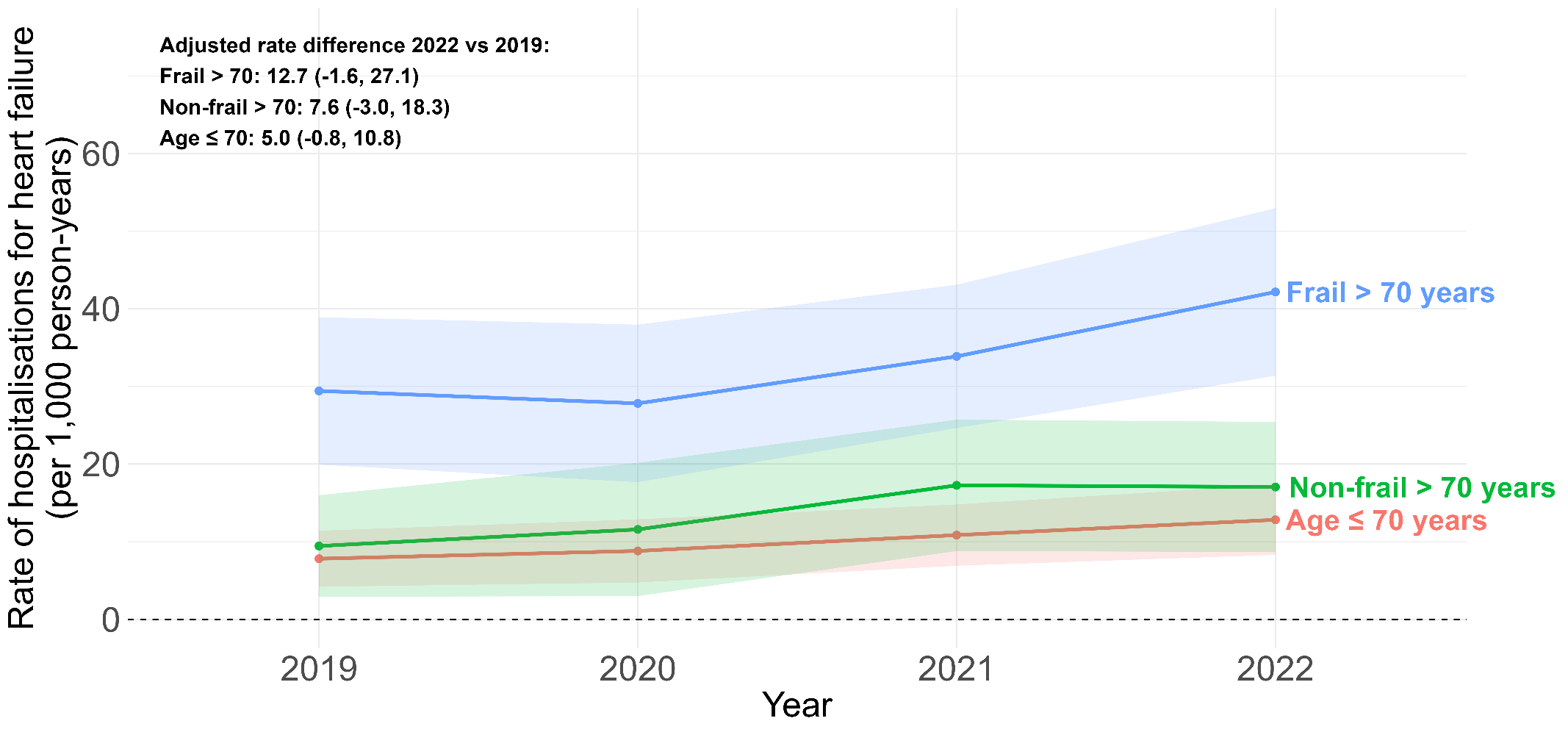

**Supplementary Figure 10.** Interrupted time series analysis of monthly SGLT2i initiations following 2022 NICE guideline update by age and frailty subgroups, 2019-2014. The solid black line shows the observed trend, the dashed red line shows the counterfactual trend, and the vertical dashed blue line shows the intervention (publication of the NICE guidelines).

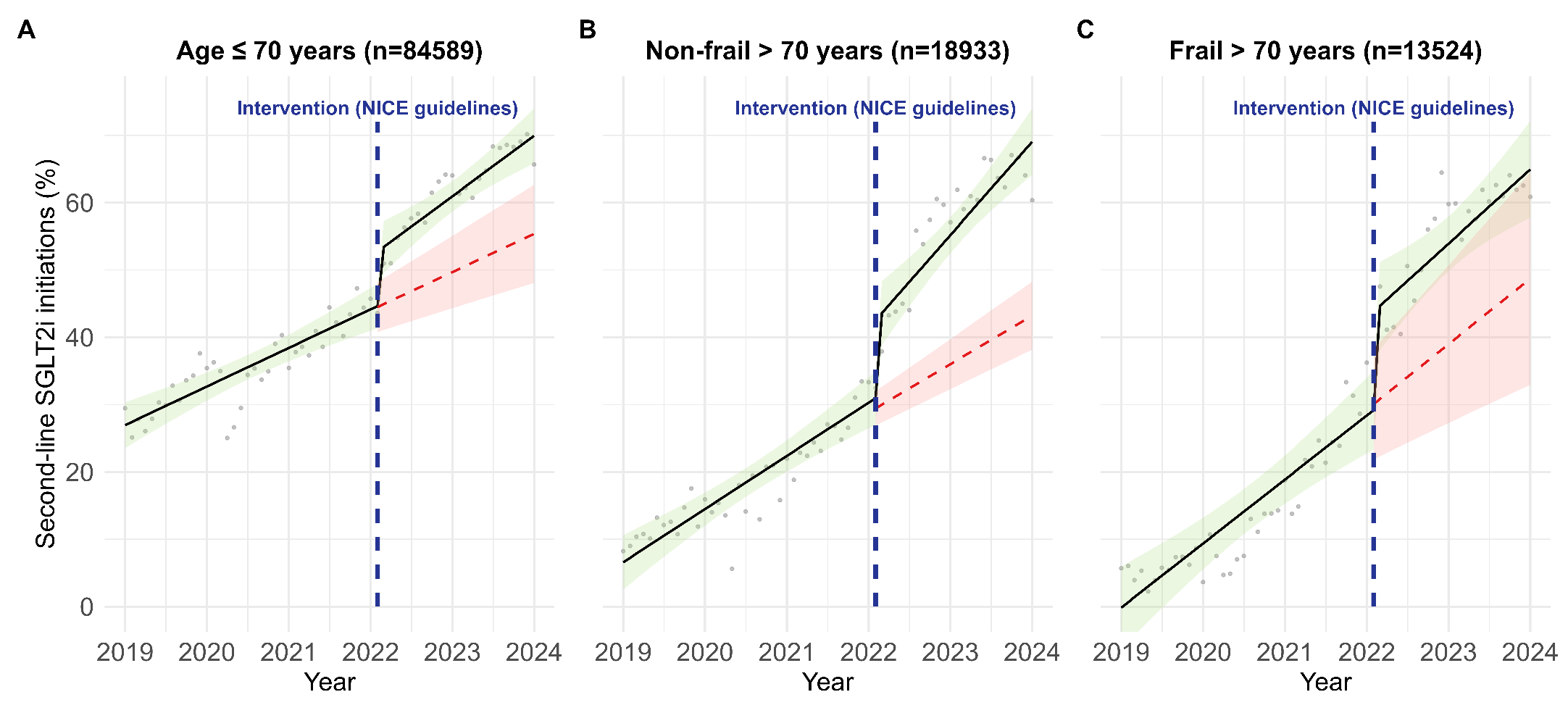
